# Volumetric and microstructural abnormalities of the amygdala in focal epilepsy with varied levels of SUDEP risk

**DOI:** 10.1101/2023.03.13.23287045

**Authors:** Antoine Legouhy, Luke A. Allen, Sjoerd B. Vos, Joana F. A. Oliveira, Michalis Kassinopoulos, Gavin P. Winston, John S. Duncan, Jennifer A. Ogren, Catherine Scott, Rajesh Kumar, Samden D. Lhatoo, Maria Thom, Louis Lemieux, Ronald M. Harper, Hui Zhang, Beate Diehl

## Abstract

Although the mechanisms of sudden unexpected death in epilepsy (SUDEP) are not yet well understood, generalised- or focal-to-bilateral tonic-clonic seizures (TCS) are a major risk factor. Previous studies highlighted alterations in structures linked to cardio-respiratory regulation; one structure, the amygdala, was enlarged in people at high risk of SUDEP and those who subsequently died. We investigated volume changes and the microstructure of the amygdala in people with epilepsy at varied risk for SUDEP since that structure can play a key role in triggering apnea and mediating blood pressure.

The study included 53 healthy subjects and 143 patients with epilepsy, the latter separated into two groups according to whether TCS occur in years before scan. We used amygdala volumetry, derived from structural MRI, and tissue microstructure, derived from diffusion MRI, to identify differences between the groups. The diffusion metrics were obtained by fitting diffusion tensor imaging (DTI) and neurite orientation dispersion and density imaging (NODDI) models. The analyses were performed at the whole amygdala level and at the scale of amygdaloid nuclei.

Patients with epilepsy showed larger amygdala volumes and lower neurite density indices (NDI) than healthy subjects; the left amygdala volumes were especially enhanced. Microstructural changes, reflected by NDI differences, were more prominent on the left side and localized in the lateral, basal, central, accessory basal and paralaminar amygdala nuclei; basolateral NDI lowering appeared bilaterally. No significant microstructural differences appeared between epilepsy patients with and without current TCS.

The central amygdala nuclei, with prominent interactions from surrounding nuclei of that structure, project to cardiovascular regions and respiratory phase switching areas of the parabrachial pons, as well as to the periaqueductal gray. Consequently, they have the potential to modify blood pressure and heart rate, and induce sustained apnea or apneusis. The findings here suggest that lowered NDI, indicative of reduced dendritic density, could reflect an impaired structural organization influencing descending inputs that modulate vital respiratory timing and drive sites and areas critical for blood pressure control.

## 1 Introduction

Although mechanisms underlying sudden unexpected death in epilepsy (SUDEP) remain unclear, monitored SUDEP cases highlighted a common pattern in which the triggering of generalised or focal-to-bilateral tonic-clonic seizures (TCS) is followed by a short period of increased heart and respiratory rates, after which a combination of central apnea, severe bradycardia and, most commonly, transient asystole occurs, accompanied by generalised postictal EEG suppression [1]. The cardiorespiratory collapse inevitably follows a tonic-clonic seizure (TCS). In case-control studies, the presence of TCS, especially at high frequency, emerges as the major risk factor for SUDEP [2]. Recent studies clarified that central apnea occurring after a TCS, so-called postictal central apnea, is a particular risk factor for SUDEP [3].

Structural MRI studies have shown alterations in several cardiorespiratory brain sites in people considered at risk for SUDEP and those who succumbed to it. Specifically, limbic structures, including the bilateral amygdala, parahippocampal gyrus, and entorhinal cortex were enlarged in SUDEP cases and high-risk subjects. The subcallosal cortex, a region involved in blood pressure regulation, was also enlarged, but in SUDEP only [4]. The amygdala is influential in triggering apnea, as shown in animal and human studies, including direct electrical stimulation of the structure [5][6]. In addition, a number of critical sites lose volume in epilepsy, in particular, sites within the cerebellum, thalamus and brainstem [4][7]. As these sites are injured, the enlarged amygdala may exert abnormal influences on cardiac and respiratory structures in a particularly vulnerable phase after a TCS. The amygdala has pronounced projections to cardiovascular regions and respiratory phase switching areas of the parabrachial pons and the periaqueductal gray. Although we showed volumetric changes in patients who are at risk for SUDEP [4][7], we lack insights into the tissue composition that lead to those changes in volume. Here, we investigate the microstructure of the amygdala in patients with and without current TCS using diffusion MR imaging. Whole amygdala and sub-nuclei were examined and compared in patients with and without TCS and healthy controls.

Diffusion MR imaging has been used to characterize brain structure at micron scales to reveal widespread changes in both focal and generalized epilepsy [8]. This technique uses MRI to interrogate the rate (diffusivity) of diffusional motion of water molecules as a function of direction. Because water diffusion is influenced by the underlying tissue microstructure, measured diffusivity can then be used to deduce important characteristics of the underlying tissue microstructure, *e*.*g*., revealing the overall direction of axons in a white matter voxel, as water diffusion is confined by myelin surrounding the fibers. The standard approach for such studies is diffusion tensor imaging (DTI); DTI can estimate in each voxel a quantity, fractional anisotropy (FA), which reflects the orientation dependence of diffusion, and infer the primary direction of the underlying axons, enabling virtual dissection of white matter bundles with tractography. Combined with the other derived quantities, such as mean diffusivity (MD), DTI can gain insights into microstructural changes for specific white matter structures of interest.

However, FA and MD can only provide non-specific characterisation of tissue microstructure. For example, a change in FA can be caused by a change in axonal density or spatial organization of axons. More advanced diffusion MR imaging techniques have been developed to address this limitation of DTI. In particular, neurite orientation dispersion and density imaging (NODDI) [9] enables differentiation of two important aspects of neuronal morphology: the packing density of neurites via the neurite density index (NDI), and the spatial organization of these neuronal projections via the orientation dispersion index (ODI); these values correlate with histology [10], [11]. NODDI, applied earlier in structural studies in epilepsy, has shown promise in delineating cortical dysplasia [12].

These microstructural and volumetric procedures have the potential to provide insights into the processes underlying the altered amygdala volumes in people with epilepsy, and the possibility that those processes may underlie the aberrant physiological consequences found after ictal events. Because functions mediated by the amygdala are asymmetrical [54], and structures with pronounced projections to the amygdala, such as the insular cortices, also exert lateralized influences [55], both left and right amygdala volumes and microstructure were considered separately and together.

## 2 Material and methods

### 2.1 Demographics

#### Study design

Participants were identified from a database of patients recruited as part of an ongoing prospective investigation into autonomic and imaging biomarkers of SUDEP (the Center for SUDEP Research; CSR) conducted at University College London (UCL) while being assessed for epilepsy surgery candidacy; age-matched healthy controls scanned at UCL were selected from a database of controls. All participants gave written informed consent (research ethics committee (19/SW/00071)).

#### Acquisition

Patients and healthy control subjects underwent high resolution anatomical and advanced diffusion imaging on the same 3T GE MR750 scanner. Anatomical acquisitions consisted of 3D T1-weighted images, voxel size 1×1×1 mm. The multi-shell diffusion acquisitions were composed of 11 b=0 and diffusion-weighted images with the following gradient properties: b-values 300, 700, 2500 *s. mm*^-2^, respective number of directions: 8, 32, 64; voxel size: 2×2×2 mm. The 0, 300 and 700 b-values are well suited for standard DTI estimation; whereas, the full protocol is optimized for NODDI.

#### Population

A total of 211 patients with focal epilepsy underwent the full imaging protocol. After visual inspection on T1-weighted and DWI images, 35 patients were excluded based on pathological criteria (large lesions, focal cortical dysplasia, tumours, stroke, severe atrophy, previous resection), and 26 were removed due to image artifacts. Finally, 7 patients were removed as they formed a cluster of older people (>54.3 years old), too different from the rest of the population.

After this outlier rejection step, a total of 143 patients with epilepsy were considered for the analysis. In addition, 53 healthy control subjects underwent precisely the same imaging protocol. We assessed a total of 196 subjects, divided into groups according to the presence and occurrence of generalized TCS:

- C: Healthy control group.
- TCSneg: Group of patients with epilepsy who never had generalized TCS, or had in the past, but not for several years.
- TCSpos: Group of patients with epilepsy who currently have occurrences of generalized TCS at least once per year, with most having more than three per year.

At the time of writing (February, 2023), 3 of the subjects included in this study subsequently died from SUDEP. Demographic details are shown in Table 1.

**Table 1:** Demographics and clinical specifications for healthy control subjects and epilepsy patients by TCS groups.

| Group | Control | TCSneg | TCSpos | All epilepsy |
| --- | --- | --- | --- | --- |
| Number of subjects | 53 | 75 | 68 | 143 |
| Age (in years)<br>mean (std) | 36.4 (10.2) | 33.3 (9.4) | 32.2 (7.6) | 32.7 (8.6) |
| Sex<br>female : male | 36 : 17 | 39 : 36 | 30 : 38 | 69 : 74 |
| Age at epilepsy onset (in<br>years) | - | 14.6 (9.1) | 15.1 (8.5) | 14.9 (8.8) |
| Disease duration (in years)<br>mean (std) | - | 18.7 (10.8) | 17.1 (9.6) | 18.0 (10.3) |
| Lesional subjects<br>number (percentage) | - | 37 (49%) | 22 (32%) | 59 (41%) |
| Epilepsy lateralization: |  |  |  |  |
| Left | - | 28 | 32 | 60 |
| Right | - | 36 | 24 | 60 |
| Non-lateralized | - | 9 | 10 | 19 |
| Unknown | - | 2 | 2 | 4 |
| Epilepsy syndrome: |  |  |  |  |
| Multifocal | - | 8 | 7 | 15 |
| Hemispheric | - | 14 | 8 | 22 |
| Temporal | - | 31 | 32 | 63 |
| Occipital | - | 0 | 2 | 2 |
| Frontal | - | 12 | 10 | 22 |
| Temporo-occipital | - | 4 | 1 | 5 |
| Fronto-temporal | - | 3 | 4 | 7 |
| Insula | - | 1 | 2 | 3 |
| Unknown | - | 2 | 2 | 4 |
| Subsequent SUDEP (follow up): |  |  |  |  |
| Definite | - | 1 | 1 | 2 |
| Probable | - | 0 | 1 | 1 |

### 2.2 Image processing

#### DWI pre-processing

DWI volumes were corrected for tissue magnetic susceptibility-induced distortion using FSL TOPUP [13][14], for signal drift using the method from [15], and simultaneously for subject motion and eddy current distortions using FSL EDDY [16].

#### Model fitting

- The DTI model was fitted through weighted linear least squares using FSL DTIFIT. At high b-values, the deviation from the Gaussian assumptions inherent in the DTI model becomes too pronounced [Jensen 2010]. We therefore only considered the volumes corresponding to b-values below 1000 *s. mm*^-2^ for this fitting. We used gradient directions corrected for subject head motion [17]. The fitting produced several scalar maps: the baseline signal image (S0), the mean diffusivity (MD) that quantifies the average mobility of water molecules in all directions, and the fractional anisotropy (FA) that quantifies diffusion anisotropy.
- The NODDI model [9] was fitted using the NODDI Matlab toolbox (http://mig.cs.ucl.ac.uk/index.php?n=Tutorial.NODDImatlab). The model decomposes the signal into a tissue compartment itself distinguishing between intra- (sticks) and extra- (cylinder) neurite space, and a free water compartment (isotropic). NODDI enables the extraction of several scalar maps, including the orientation dispersion index (ODI) that quantifies the angular variation of neurites reflecting the spatial organization of the neuronal projections, the neurite density index (NDI) that corresponds to the fraction of intra-neurite signal among the tissue compartment appraising the packing density of neurites, and the free water volume fraction (FWF) that measures the extent of CSF contamination.

#### Segmentation

Whole-amygdalae segmentations were performed on T1-weighted images using Freesurfer software (version 7.0.0) [18][19]. The amygdalae were then subdivided using the method developed in [20], which uses a segmented atlas from manual delineations on post-mortem subjects who have undergone specific very high-resolution anatomical acquisition to distinguish the amygdala nuclei. After removing regions that were smaller than the diffusion image resolution, 5 amygdaloid labels remained on each side of the brain: lateral nucleus, basal nucleus, central nucleus, accessory basal nucleus and paralaminar nucleus. An example of the output segmentations on a subject is shown in Fig. 1.

**Figure 1:**
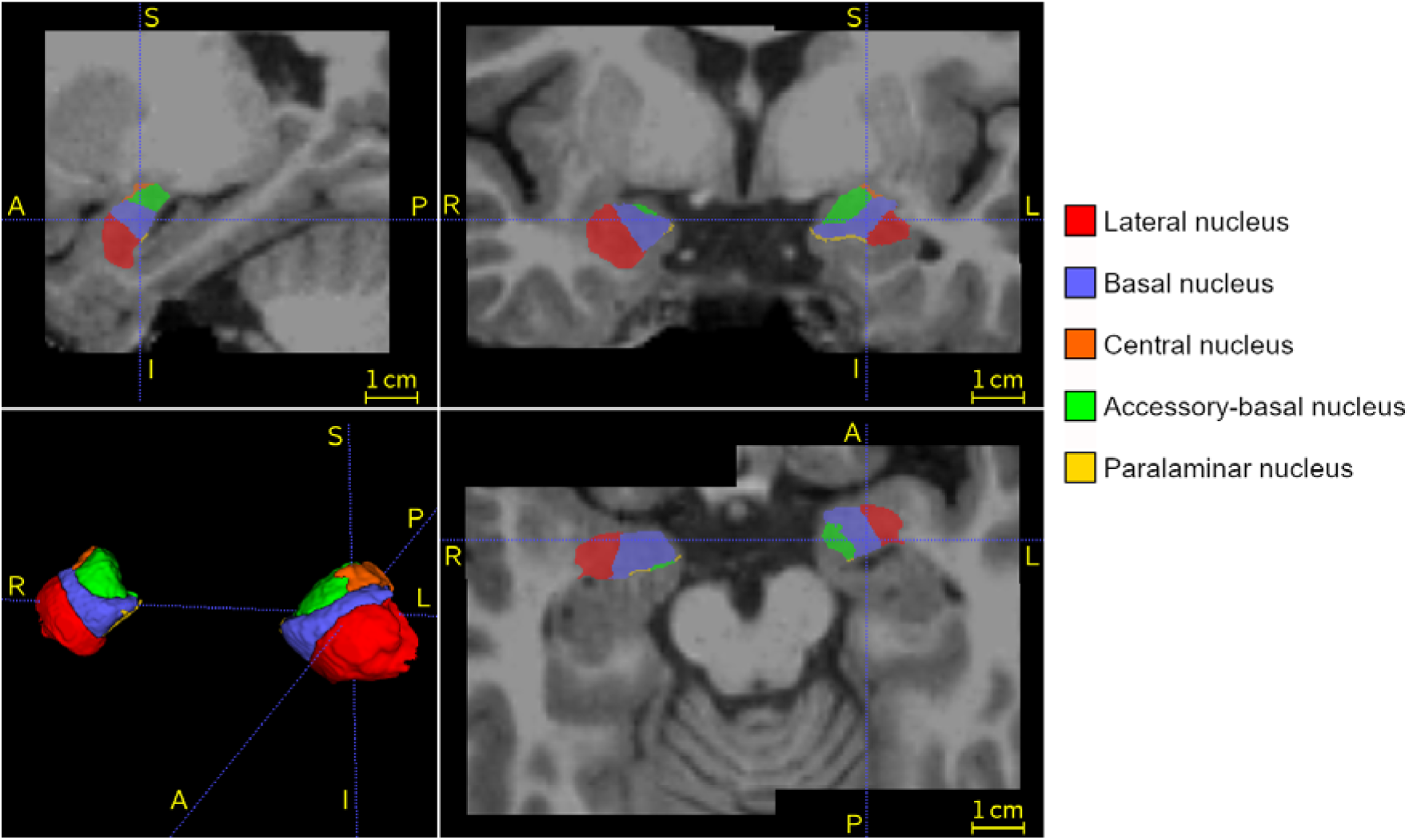
Amygdaloid nuclei segmentation in a representative patient.

#### Volume extraction

We used the surface-based volumetric statistics generated by Freesurfer to assess the volumes of the segmented amygdaloid structures. Those volumes were then normalized by dividing by that of the whole brain.

#### Regional diffusion metric extraction

Prior to the extraction of regional statistics of diffusion metrics, diffusion scalar maps were mapped to the segmentation space through an affine registration of the S0 image onto the T1-weighted image using Anima software [https://anima.irisa.fr]. To minimise the effect of CSF partial volume, we chose to capitalize on the free water fraction (FWF) estimated in the NODDI fitting to compute tissue-weighted means following [21]. Given a metric map of interest M and a tissue fraction map TF=1-FWF, the tissue-weighted mean 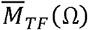 of the voxels contained in the ROI Ω is computed as follows:

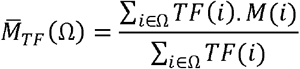

### 2.3 Statistical analysis design

Multivariate analysis of covariance (MANCOVA) was used to assess, for each amygdaloid region, whether differences emerged in diffusion metrics between the GTCS groups after correcting for the influence of age and sex. The dependent variables were the ROI-wise tissue-weighted means of the diffusion metrics for each subject; the null hypothesis (H0) was that the means across groups were equal for each dependent variable; the considered multivariate statistic is the Lawley-Hotelling trace (although Wilks’ lambda or Pillai’s trace would have given similar results in our context) [53]. The analyses were performed for volumes and then separately for DTI and NODDI metrics and at two scales: whole amygdala structure and amygdaloid subfields.

In cases of significance of the multivariate test, analysis of covariance (ANCOVA) was performed for each dependent variable individually, also with correction for age and sex.

In case of significance of this univariate test, subsequent post-hoc t-tests were used for exhaustive comparison between groups (TCSpos vs control, TCSneg vs control, TCSpos vs TCSneg), also correcting for age and sex.

For each family of tests, the Bonferroni correction with 5% FWER was performed to correct for multiple comparisons. For each t-test we reported the p-value *p* and Cohen’s d effect size *d*.

## 3 Results

### 3.1 Whole amygdala

We first compared the volumes and diffusion metrics of left and right amygdalae taken as whole structures between epilepsy groups.

#### 3.1.1 Volume

Bilateral amygdala volumes were significantly larger in patients with ongoing TCS compared to the healthy control subjects; (*p*<0.001) with a medium effect size (*d*>0.6). Comparing patients with epilepsy with current TCS against those without, the left amygdala was significantly larger in the TCSpos group (*p*<0.01), with a medium effect size (*d*=0.5). A similar trend appeared in the right side but did not reach significance. Results are detailed in Table 2; associated box and violin plots are in Fig. 2.

**Table 2:** Group comparison statistics for normalized volumes, DTI metrics (MD, FA) and NODDI metrics (ODI, NDI). The displayed p-values correspond to the pairwise t-tests (which were performed only if the prior ANCOVA was significant), they were uncorrected. FWER correction with 5% FWER: 6 t-tests for volumes, 3 for DTI metrics and 9 for NODDI metrics. Cohen’s d were computed following: group on the left minus group on the right (e.g. for C vs TCSneg: —————————. Significant P values are framed red.

| Amygdala |  |  |  |  |  |  |  |  |  |  |
| --- | --- | --- | --- | --- | --- | --- | --- | --- | --- | --- |
| left |  |  |  |  |  |  |  |  |  |  |
| group | normalized volume ($\times 10^{-3}$) | | MD ($\mu\text{m}^2/\text{ms}$) | | FA | | ODI | | NDI | |
|  | mean | std | mean | std | mean | std | mean | std | mean | std |
| C | 1.394 | 0.159 | 0.850 | 0.031 | 0.237 | 0.013 | 0.425 | 0.019 | 0.444 | 0.015 |
| TCSneg | 1.435 | 0.162 | 0.871 | 0.035 | 0.235 | 0.016 | 0.408 | 0.022 | 0.423 | 0.023 |
| TCSpos | 1.530 | 0.203 | 0.868 | 0.027 | 0.230 | 0.015 | 0.416 | 0.022 | 0.421 | 0.019 |
|  | p-value | Cohen's d | p-value | Cohen's d | p-value | Cohen's d | p-value | Cohen's d | p-value | Cohen's d |
| C vs TCSneg | 2.80E-01 | -0.171 | 1.90E-03 | -0.595 | - | 0.104 | 3.40E-05 | 0.721 | 7.20E-08 | 0.945 |
| C vs TCSpos | 5.10E-04 | -0.669 | 3.80E-03 | -0.476 | - | 0.434 | 7.00E-02 | 0.309 | 4.70E-10 | 0.990 |
| TCSneg vs TCSpos | 3.20E-03 | -0.498 | 4.70E-01 | 0.119 | - | 0.330 | 1.60E-02 | -0.412 | 7.80E-01 | 0.046 |

Table 2: Group comparison statistics for normalized volumes, DTI metrics (MD, FA) and NODDI metrics (ODI, NDI).
| right |  |  |  |  |  |  |  |  |  |  |
| --- | --- | --- | --- | --- | --- | --- | --- | --- | --- | --- |
| group | normalized volume ($\times 10^{-3}$) | | MD ($\mu\text{m}^2/\text{ms}$) | | FA | | ODI | | NDI | |
|  | mean | std | mean | std | mean | std | mean | std | mean | std |
| C | 1.597 | 0.158 | 0.830 | 0.027 | 0.244 | 0.020 | 0.428 | 0.019 | 0.448 | 0.016 |
| TCSneg | 1.676 | 0.159 | 0.841 | 0.044 | 0.242 | 0.025 | 0.417 | 0.026 | 0.432 | 0.029 |
| TCSpos | 1.715 | 0.234 | 0.847 | 0.052 | 0.238 | 0.019 | 0.422 | 0.026 | 0.429 | 0.031 |
|  | p-value | Cohen's d | p-value | Cohen's d | p-value | Cohen's d | p-value | Cohen's d | p-value | Cohen's d |
| C vs TCSneg | 8.60E-03 | -0.392 | - | -0.314 | - | 0.175 | - | 0.403 | 2.60E-04 | 0.582 |
| C vs TCSpos | 3.20E-03 | -0.587 | - | -0.485 | - | 0.453 | - | 0.163 | 2.90E-05 | 0.722 |
| TCSneg vs TCSpos | 2.60E-01 | -0.196 | - | -0.171 | - | 0.278 | - | -0.239 | 4.40E-01 | 0.139 |

**Figure 2:**
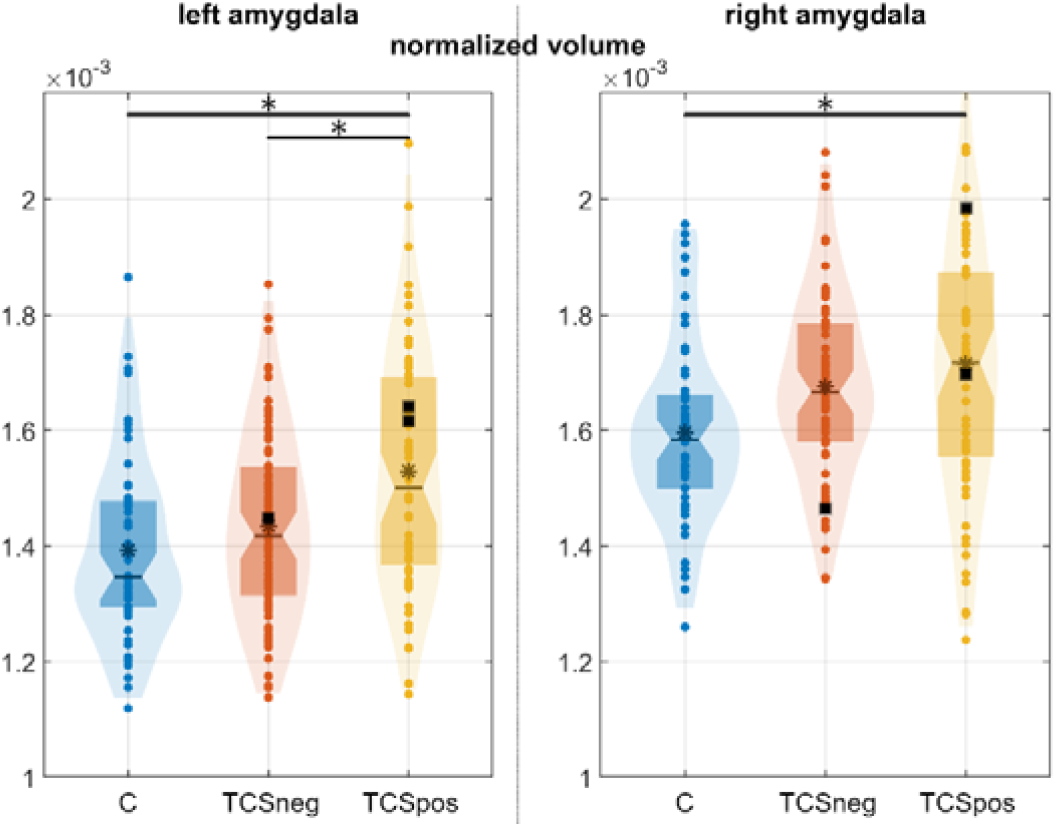
Box and violin plots of the normalized volume in the amygdalae. ⍰ indicates significant difference between groups for 5% FWER. Black squares represent the subjects who subsequently died from SUDEP.

#### 3.1.2 DTI metrics

Compared to healthy controls, higher MD appeared in the left amygdala in patients with epilepsy (C vs TCSneg (*p*<0.01) with medium effect size (*d*=0.6); C vs TCSpos (*p*<0.01), medium effect size (*d*=0.5); see Table 2 and Fig. 3. A trend of reduced FA in both epilepsy groups compared to controls was not significant.

**Figure 3:**
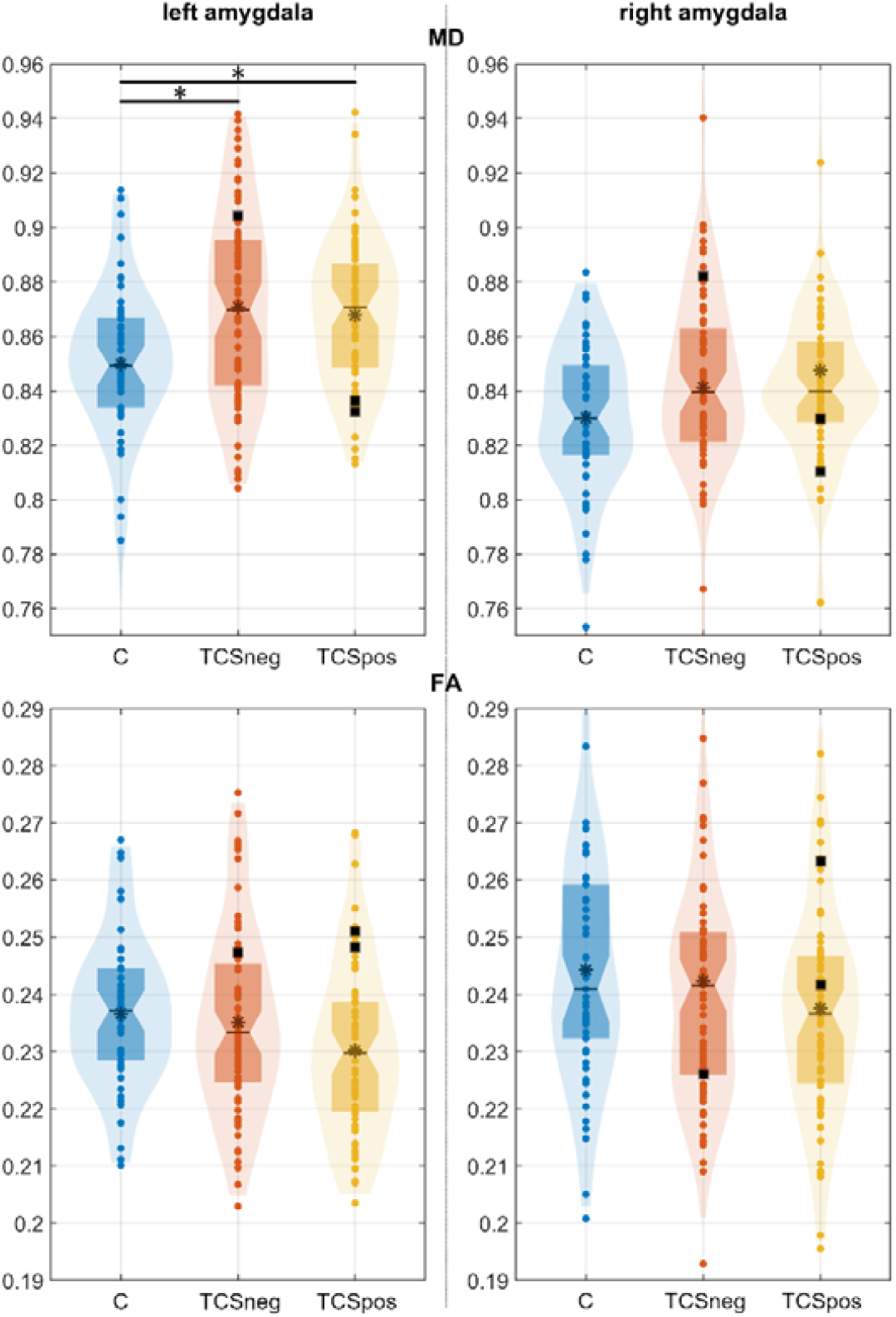
Box and violin plots of the MD (in μm^2^ / ms) and FA metrics in the amygdalae. ⍰ indicates significant difference between groups for 5% FWER. Black squares represent the subjects who subsequently died from SUDEP.

A bilateral trend of patients with epilepsy with current TCS (TCSpos) showing higher MD and lower FA in the amygdala compared to patients with epilepsy currently without (TCSneg) was not significant.

#### 3.1.3 NODDI metrics

Compared to healthy controls, NDI in left and right amygdala was lower in both epilepsy groups.

C vs TCSneg (left: *p*<10e-7 with large effect size: *d*>0.9; right: *p*<0.001 with medium effect size: *d*=0.6); C vs TCSpos (left: *p*<10e-9 with large effect size: *d*=1; right *p*<10e-4 with quite large effect size on the right: *d*=0.7); see Table 2 and Fig. 4. There was no difference between epilepsy groups, although patients with epilepsy with current TCS (TCSpos) showed nominally slightly lower NDI in the left and right amygdala compared to epilepsy patients without TCS (TCSneg).

**Figure 4:**
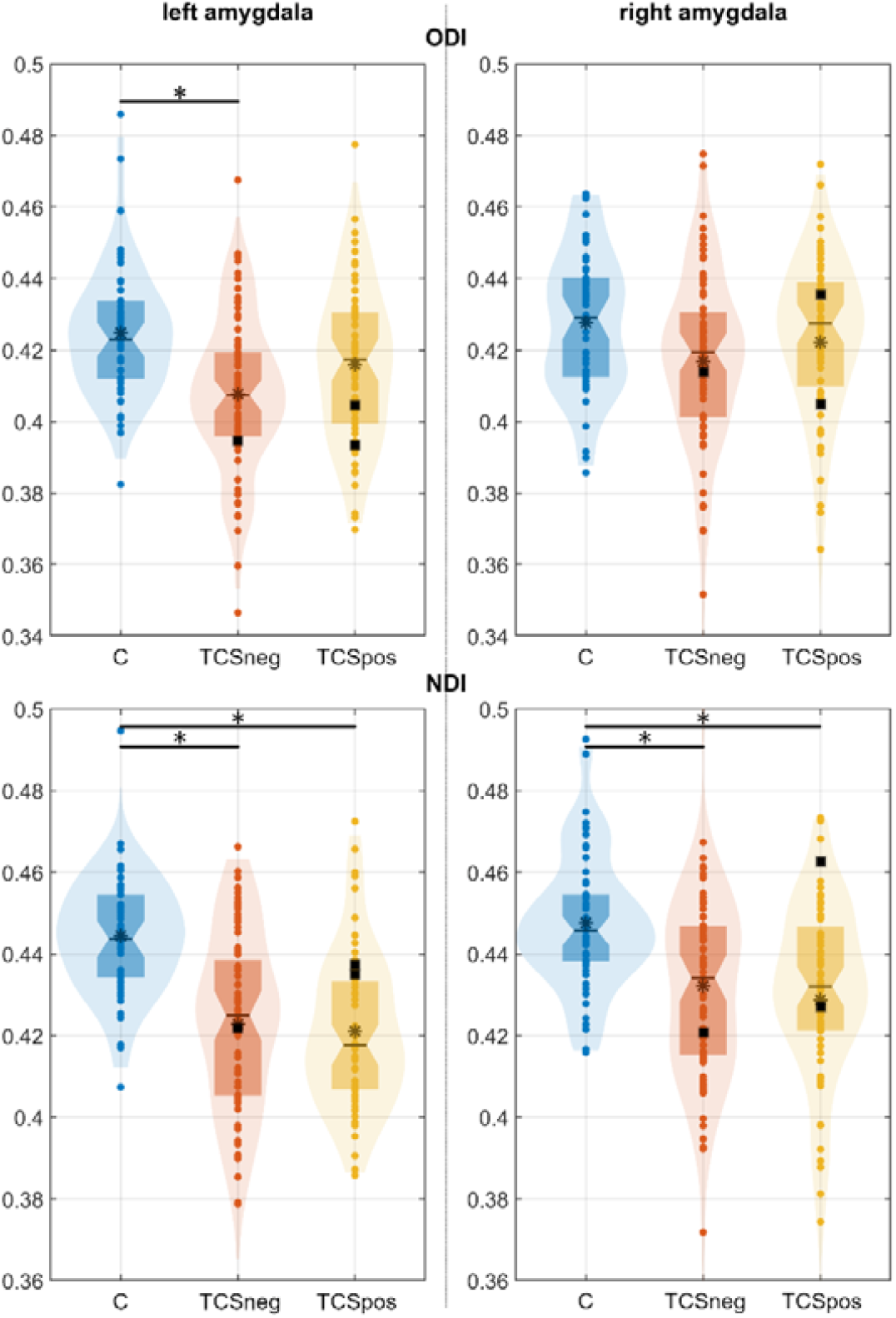
Box and violin plots of the ODI and NDI metrics in the amygdalae. ⍰ indicates significant difference between groups for 5% FWER. Black squares represent the subjects who subsequently died from SUDEP.

ODI metrics showed a trend to be bilaterally lower in TCSneg and TCSpos group compared to the C groups. Surprisingly, the values were even lower in the TCSneg group. The differences in ODI were significant only on the left side for C vs TCSneg (*p*<0.0001) with a large effect size (*d*=0.7).

### 3.2 Amygdala nuclei

In general, volumetric and diffusion measurements of the amygdala nuclei followed the same trends as the whole amygdala. Table 3 of the Appendix lists all interrogated nuclei.

**Table 3:**
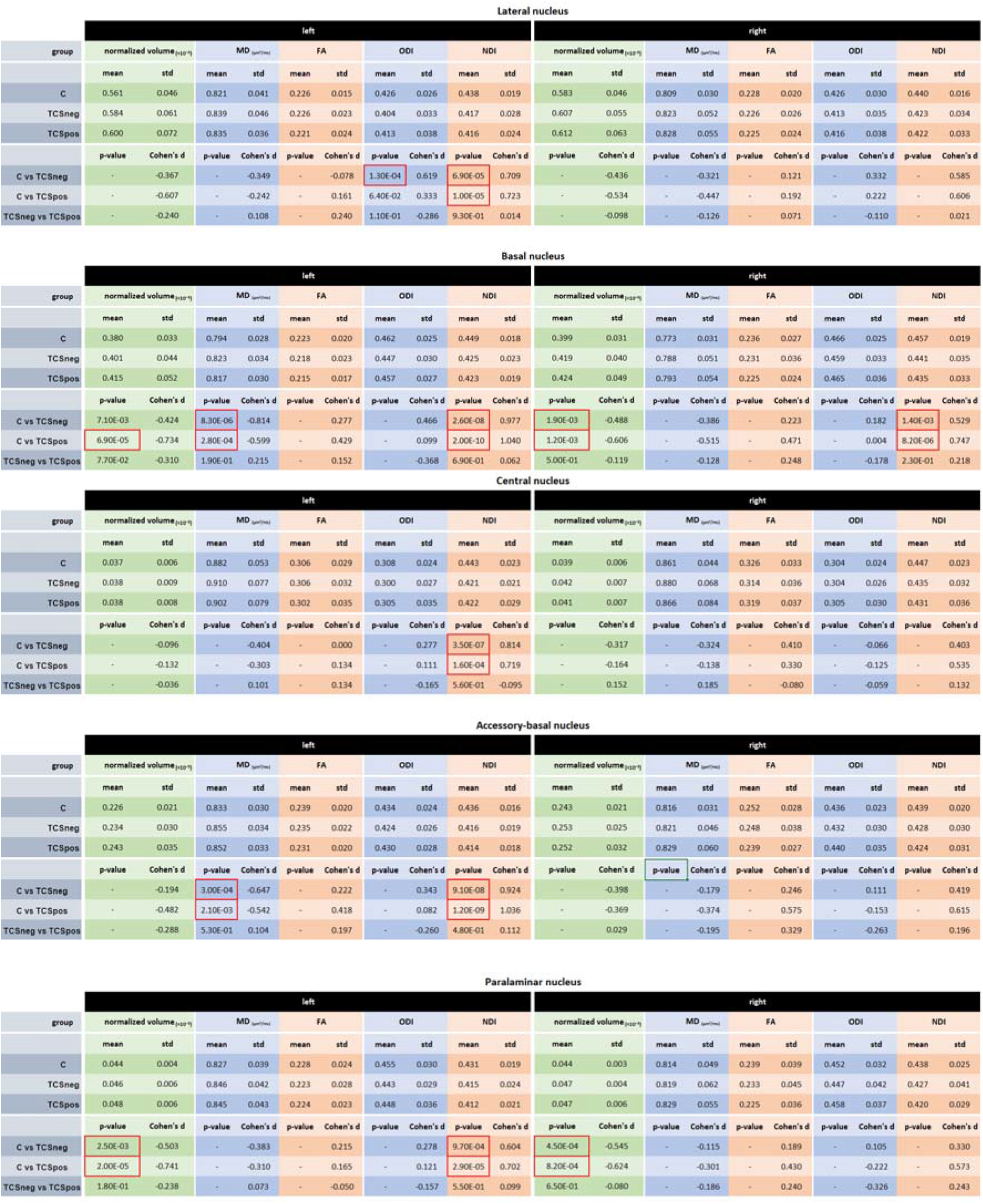
Group comparison statistics for normalized volumes, DTI metrics (MD, FA) and NODDI metrics (ODI, NDI) in amygdala nuclei. The displayed p-values correspond to the pairwise t-tests (which are performed only if the prior ANCOVA is significant), they are uncorrected. FWER correction with 5% FWER: 6 t-tests for volumes, 3 for DTI metrics and 9 for NODDI metrics. Cohen’s d are computed following: group on the left minus group on the right (e.g. for C vs TCSneg: 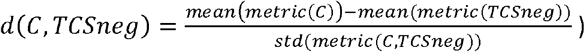.

The nominal volumes of the individual amygdala nuclei were largest in patients with epilepsy with current TCS, followed by those without current TCS, and volumes were smallest in healthy controls. The diffusion tensor metrics reveal nominally highest mean diffusivity in TCSpos, and healthy controls lowest. Fractional anisotropy was highest in controls, and lowest in the TCSpos group.

NODDI metrics revealed highest neurite density index (NDI) in controls, and these values were lower in TCSpos compared to TCSneg; the orientation dispersion index was also highest in controls. Only the significant differences are reported in the following.

#### 3.2.1 Volume

##### Basal nucleus

There were significantly larger volumes bilaterally in the TCSpos group compared to C (left: *p*<0.0001, right: *p*<0.01) with large effect sizes (left: *d*>0.7, right: *d*>0.6). On the right side, volumes were also larger in the TCSneg group compared to C (*p*<0.01) with a medium effect size (d=0.5).

##### Paralaminar nucleush

There were significantly larger volumes bilaterally in the TCSneg group compared to C (left: *p*<0.01, right: *p*<0.001), with medium effect sizes (left: d>0.5, right: d>0.5), and in the TCSpos group compared to C (left: *p*<0.0001, right: *p*<0.0001) with quite large effect sizes (left: d>0.7, right: d>0.6).

All other nuclei examined showed the above-described nominal variation, but differences were not significant.

#### 3.2.2 SUDEP victims

The small number of SUDEP cases (n=3), and the mixed outcomes of values on laterality, volume, and microstructure precluded generalizations of those measures to the fatal event. On inspection, values tended to be near-outliers on most measures of volume, MD, FA, ODI and NDI, but laterality, direction of change, and TCS history varied to the extent that summary profiles would be premature.

#### 3.2.3 DTI metrics

##### Basal nucleus

Significantly higher MD values appeared on the left side in the TCSneg group compared to C (*p*<10e-5), with a large effect size (d>0.8), and in the TCSpos group compared to C (*p*<0.001) with a medium effect size (d=0.6).

##### Accessory-basal nucleus

There were significantly higher MD on the left side in the TCSneg group compared to C (*p*<0.001) with a medium effect size (*d*>0.6), and in the TCSpos group compared to C (*p*<0.01) with a medium effect size (*d*>0.5).

#### 3.2.4 NODDI metrics

##### Lateral nucleus

There were significantly lower NDI on the left side in the TCSneg group compared to C (*p*<0.0001) with a quite large effect size (*d*>0.7), and in the TCSpos group compared to C (*p*=10e-5) with a quite large effect size (*d*=0.6). There were significantly lower ODI on the left side in the TCSneg group compared to C (*p*<0.001) with a medium effect size (*d*>0.6).

##### Basal nucleus

Significantly lower NDI bilaterally appeared in the TCSneg group compared to C (left: *p*<10e-7, right: *p*<0.01) with a large effect size on the left, medium on the right (left: *d*=1, right: *d*>0.5), and in the TCSpos group compared to C (left: *p*<10e-9, right: *p*<10e-5) with a large effect size on the left, and quite large on the right (left: *d*>1, right: *d*>0.7).

##### Central nucleus

There were significantly lower NDI on the left side in the TCSneg group compared to C (*p*<10e-6) with a large effect size (*d*>0.8), and in the TCSpos group compared to C (*p*<0.001) with a quite large effect size (*d*=0.7).

##### Accessory-basal nucleus

There were significantly lower NDI on the left side in the TCSneg group compared to C (*p*<10e-7) with a large effect size (*d*>0.9), and in the TCSpos group compared to C (*p*<10e-8) with a large effect size (*d*>1).

##### Paralaminar nucleus

Significantly lower NDI appeared on the left side in the TCSneg group compared to C (*p*<0.001) with a medium effect size (*d*>0.6), and in the TCSpos group compared to C (*p*<0.0001) with a quite large effect size (*d*>0.7).

The results are summarized in Table 3.

## 5 Discussion

We evaluated volumes and microstructure of the amygdala using standard structural and advanced diffusion MR imaging procedures respectively in patients with epilepsy compared to healthy controls, and revealed volume changes accompanied by significant microstructural alterations. Subgroup analyses of patients with epilepsy, grouped based on their occurrences of TCS, revealed that patients with epilepsy with current TCS overall have larger amygdala volumes than healthy controls. Within the epilepsy groups, patients with current occurrences of TCS also had larger left amygdalae than those who did not. The microstructural analyses, using diffusion imaging, revealed increased diffusivity in the left amygdala, and no significant FA changes. Amongst NODDI parameters, NDI appeared most sensitive to reveal changes, with bilateral reductions in the amygdala in both epilepsy groups relative to controls.

### Amygdala and Volume changes

We earlier found increased amygdala volumes for patients with more than three GTCS per year compared to healthy controls [4]; these increases appeared whether using VBM or regional segmentation. Increased bilateral amygdala and hippocampal volumes appear in a subtype of subjects with mesial temporal lobe epilepsy [22]; these subjects also have the poorest surgical outcome and are at greatest risk of SUDEP [23].

A recent review identified 107 patients in eight papers providing reasonable evidence that in temporal lobe cases, mesial temporal structures, including the amygdala, were involved in seizure onset [24]. In some cases, reduction of amygdala volume was accompanied by improved (lowered) seizure frequency and treatment [25]. Overall, seizure frequency was higher in patients with focal unaware seizures with amygdala enlargement, and TCS, although present, comprised a lower percentage of their seizures [24]. Notably, greater seizure frequency appeared in people with larger amygdalae, and antiseizure medication was associated with reduced amygdala volumes.

The broader literature on amygdala volume relationships suffers from inhomogeneous groups and difficulties in ascertaining amygdala enlargement. Of interest, in one study, people with larger amygdalae had a greater likelihood of postictal psychosis [26][27]. A larger percentage showed depression and anxiety, implicating a relation between behavioral alterations and structural alterations in the amygdalae.

Patients included in our series all had focal epilepsy; the majority had temporal lobe epilepsy, but the cohort was inhomogeneous with respect to the epilepsy syndrome. A large group had frontal lobe epilepsy, and in many, the seizure focus could only be lateralized or localized to the anterior or posterior quadrants. Therefore, seizure onset may originate from multiple sites other than the amygdala, but the structure receives substantial input from other sites involved in cardiorespiratory regulation.

Demographic variables, including age, epilepsy duration and age of epilepsy onset were similar in both our epilepsy groups, and should not contribute major influences on these results. The control and epilepsy groups were of similar age. Female-to-male ratios were comparable in both epilepsy groups; however, the number of female participants in the control group was twice that of males.

With all groups combined, we observed volumes about 14% larger in the right amygdala compared to the left. This asymmetry has been documented in a group of healthy control subjects [29] or [30], but the literature is inconsistent in that aspect; a meta-analysis can be found in [31]. Rather than an anatomic feature, this rightward asymmetry may be caused by a software bias as [32] showed that the asymmetry appears with Freesurfer, but not with FIRST or manual segmentation. However, our analysis is agnostic to this aspect, since we did not perform left versus right comparisons.

The relationship between seizure types and amygdala enlargement remains understudied, as assessed by surgical resection techniques, a possible consequence of surgical logistics. Such procedures allow only for limited analysis of amygdala tissue due to the small size and often fragmented nature of the structure. During surgical resection, some nuclei in the dorsal medial aspect of the amygdala are often not removed due to their proximity to the optic tract, cerebral peduncles, and important basal forebrain regions [28]. Inferring consequences of tissue enlargement from seizure types is thus, difficult, and provided an impetus for the NODDI studies described here.

### Microstructural alterations underlying volume changes

Volume increases may result from multiple processes, including enhanced activation from experiences and learning [33][34], inflammatory processes [35]. Underlying mechanisms for gray matter changes observed on imaging may involve neurogenesis and alterations in neuronal morphology, vascular changes, and gliogenesis [33], specifically astrocytic edema potentially followed by gliosis [56]. The current study, using advanced diffusion imaging procedures, allows the potential to gain insights into the underlying microstructural changes, and differentiate the various possibilities.

The standard diffusion model, diffusion tensor imaging (DTI), allows the estimation of fractional anisotropy (FA) which reflects the orientation dependence of diffusion, infers the primary direction of the underlying axons, and assesses diffusivity.

In this study, FA and mean diffusivity were less sensitive to changes in the amygdala than NODDI metrics, which do not have the same issues as DTI, such as inappropriate assumptions of Gaussian distribution of water motion. NODDI, in providing estimates of neurite density index (NDI) and orientation dispersion index (ODI) [36], can supply insights that correlate with histological changes, as has been demonstrated in studies of the corpus callosum in both healthy brain [37] and in pathological cases, such as demyelinating spinal lesions. As a biophysical model, NODDI has shown relevance in clinical studies by being sensitive to disease-related microstructure alterations and offering biologically plausible interpretations of DWI data [38].

Recently, NODDI was used to characterize age associations with microstructural properties of amygdala subnuclei and amygdala-related white matter connections across adolescence (N = 61, 26 males; ages of 8–22 years) [39]. Age-related increases in the NDI in the lateral nucleus (LA), dorsal and intermediate divisions of the basolateral nucleus (BLDI), and ventral divisions of the basolateral and paralaminar nuclei appeared. Thus, the procedure has been used to assess neurite architecture during developmental periods of interest to those in the epilepsy field.

Postmortem studies have shown evidence of microglial activation in SUDEP, including in some cortical and subcortical regions with known autonomic functions, such as the thalamus and superior temporal gyrus. These insights may be relevant to cellular pathomechanisms underlying cardioregulatory failure during a seizure which involves central autonomic regions. The amygdala was not examined in that study [40]. In addition, microglial density was associated with cortical thinning from the ENIGMA study in some, but not all regions [41], and neuronal hypertrophy appeared in some cortical regions in SUDEP post-mortem which may potentially influence local tissue volumes. Specific studies of the amygdala in SUDEP to date did not identify increased HLA- DR- positive microglia relative to that in epilepsy and non-epilepsy control groups using a semi-quantitative scoring method [42].

Amygdala gliosis is a common finding in temporal lobe epilepsy [43]. Histological changes of amygdala nuclei, and in particular, the central nuclei, were examined in a post-mortem study in

SUDEP cases, given the known projections of the central nuclei of the amygdala to cardiac and respiratory brainstem nuclei [43]. Lower neuronal densities and increased astrocytic densities were noted in the lateral nucleus of amygdala, but not the central nucleus, compared to control tissue. The limitation of the study by Thom et al was the lack of epilepsy controls. However, assuming that the patients with the lowest NDI also have uncontrolled TCS, suggesting they are at higher SUDEP risk, the reduced NDI observed here may be an expression of such changes. Reduced neuropetidergic neuronal cell and processes (NPY, galaninergic and somatostatin) were noted in SUDEP post-mortem cases compared to the control group (epilepsy and non-epilepsy) [44]. More recently, increased amygdala serotonergic axonal networks in post-mortem SUDEP compared to epilepsy control groups were shown, and were particularly enriched in the paralaminar nucleus, but with higher density of serotonergic neurones in the basal and accessory basal nuclei of the amygdala and peri-amygdala cortex in SUDEP [45]. These findings suggest that specific alterations of neuronal populations and axonal networks in the amygdala in epilepsy and SUDEP occur, and may be reversible, but could influence NODDI metrics.

The pathological basis of amygdala hypertrophy has also been investigated in surgical series. Enlargement of the amygdala on MRI has been increasingly recognised, particularly in otherwise ‘MRI-negative’ mTLE. A systematic review of 361 cases noted that imaging criteria for amygdala enlargement remained to be clearly defined, as well as heterogeneity of the underling pathology, which included reports of astrogliosis, oligodendrogliosis, dysplasia (neuronal enlargement), low grade tumours and autoimmune-related encephalitis [46].

From this study, NDI was the diffusion metric that better differentiated the epilepsy groups. Widespread reductions in NDI occurred in patients with epilepsy in the left amygdala, involving the lateral, basal, central, accessory basal and paralaminar nuclei; the only significant change in the right amygdala involved a reduction in NDI in the basal nucleus. Of other metrics, only mean diffusivity showed increases in the left amygdala in the basal and accessory basal nucleus in all patients with epilepsy compared to controls. The above-mentioned cell enlargement is consistent with the observed NDI decrease and MD increase.

The distorted microarchitecture of the amygdala should be seen in the context of the influence the nuclei within that structure exert on breathing and cardiovascular control. Single-pulse stimulation of the central nucleus will pace inspiratory efforts in animal models, an effect that is sleep-state dependent [47]. Trains of electrical stimulation in the medial aspect of the amygdala induced apnea in children [6]. Using a machine learning algorithm, a site was identified which overlaps the very medial aspect of the basal nuclei and the cortical and medial nuclei (BL/BM, CMN). This finding, as in a previous study [48], did not depend on laterality. In this study, NDI was most prominently reduced in the bilateral basal nucleus relative to healthy controls in both TCSneg and TCSpos groups. Of interest, the basal nucleus also showed an increase in volume in epilepsy compared to controls.

The most prominent means for the amygdala to influence both cardiovascular action and respiration rests with the major projections to the periaqueductal gray (PAG) and parabrachial pons from the amygdala central nucleus [49]. The parabrachial pons serves as a respiratory phase-switching area (from inspiration to expiration and vice versa) and can thus mediate onset of apnea (by delaying inspiratory efforts), or by sustaining inspiration (apneusis). The PAG is also targeted by amygdala central nucleus projections; the PAG supports respiratory efforts [50] and appears prominently in volume changes in patients at risk for SUDEP [4]. The PAG and parabrachial pons also serve significant roles in control of blood pressure via the amygdala central nucleus and amygdala interactions with the insular and cingulate cortex. Subregions within the amygdala also influence blood pressure as well; chemical inhibition of the basal medial amygdala nucleus in rats by microinjection of muscimol (γ-aminobutyric acid (GABAA) agonist) promoted increases in mean arterial pressure (MAP) and heart rate (HR) [51]. Since loss of blood pressure is a significant consideration in processes leading to sudden death, and observed SUDEP events typically include sustained apnea [52], alterations in the amygdala that influence blood pressure and breathing in the high-risk epilepsy groups are a concern.

Although both the left and right amygdala showed volumetric and microstructural changes in patients with epilepsy that may pose risk for SUDEP based on the key role for that structure in modulating cardiovascular and breathing patterns, the findings here suggest that the alterations in the left amygdala may exacerbate the potential to disrupt those patterns, as we earlier suggested from functional connectivity data [57]. NDI was preferentially reduced in the left central nucleus between epilepsy patients compared to controls, as well as in the lateral, accessory basal and paralaminar nuclei, although lowered bilaterally in the basal amygdala nuclei. Although overall amygdala volumes were increased in people with epilepsy compared to those in healthy controls, and right whole-amygdala volumes were larger than left, the left volumes were especially elevated over the left amygdala volumes in healthy controls in our two epilepsy groups. In particular, left amygdala volumes were significantly larger in participants with TCS. Considering the projections (and the parasympathetic influences) of the left insula [55] on the amygdala, the more-affected left amygdala may be inducing deficits in parasympathetic influences on blood pressure, leading to hypotension at a critical time. A breakdown in the organization of neuronal structure can lead to unpredictable influences on nuclear output to brainstem structures; hyperactive or uncontrolled output might be expected.

The alterations found here may result from multiple influences, including excitotoxic injury, with inflammatory processes especially contributing to increased amygdala volumes. The underlying histological changes and their correlation with dysfunction of respiratory central regulation will require further studies.

## 4 Conclusion

Larger amygdala volumes in patients with epilepsy are accompanied by reduced neurite density in amygdala nuclei that project to downstream cardiovascular and respiratory regulatory sites. Such a disruption in nuclear organization likely contributes to impaired control of blood pressure and breathing, and thus, enhances risk for sudden death. The increased volumes appear to reflect a breakdown in organization rather than enhanced regulatory control.

## Data Availability

Data is not available by default.
Although, one might contact the authors for request.

## 5 Acknowledgments

This work has been supported by the National Institute of Neurological Disorders and Stroke, Grant/Award Number: U01-NS090407 (SL, BD, RH, AL), NINDS – NS090405 (SL). GPW was supported by the Medical Research Council (G0802012, MR/M00841X/1). Support was also provided by the National Institute for Health Research and University College London Hospitals Biomedical Research Centre.

